# Adventurousness cuts both ways: a Mendelian randomization of adventurousness on 10 cognitive and behavioral traits

**DOI:** 10.1101/2020.03.12.20034918

**Authors:** Charleen D. Adams, Brian B. Boutwell

**Affiliations:** Beckman Research Institute, City of Hope National Medical Center, Duarte, California, USA; School of Applied Science, The University of Mississippi P.O. Box 1848, University, MS, 38677, USA; John D. Bower School of Population Health, University of Mississippi Medical Center, Jackson, MS, 39216, USA

**Keywords:** adventurousness, risky behavior, cognitive traits, education, intelligence, Mendelian randomization

## Abstract

Adventurousness is speculated to improve happiness but also predispose to risky behaviors. Since non-experimental studies can suffer from confounding and reverse causation, and personality traits cannot be randomized, it is challenging to unravel how adventurousness impacts the mind and behavior. Mendelian randomization (MR), a quasi-randomization technique that uses genetic variants as instruments to avoid confounding and reverse causation, is an attractive option in this setting. We used MR to explore self-reported adventurousness and 10 cognitive and behavioral traits. Adventurousness decreased neuroticism and mood swings and increased years of schooling. In contrast, it also predisposed to risky behaviors (increasing the number of lifetime sexual partners, the propensity to speed in an automobile, and lifetime smoking, and decreasing the age of first having sexual intercourse). The results suggest being adventurous “cuts both ways”, evincing bivalent influences and underscoring the reality that trade-offs often accompany many human personality constructs.

---

“Adventure is not outside man; it is within”—George Eliot

The English novelist and maven of the mind, George Eliot, if the quote is not apocryphally attributed to her^1^, had grasped something important about adventure seeking more than a century before psychologists formalized the phenotype or found any genetic variants associated with it. Agnostic as to whether this “innateness” is good or bad, “adventure” seems appealing given that it conjures *both* excitement and hazard. Several of the definitions in the American Heritage Dictionary, in fact, list “exciting” and “hazardous” to describe this bivalent trait.

One may indeed speculate widely, moreover, about the usefulness of this personality construct. A sense of adventure, broadly defined, may have aided knowledge acquisition in the Enlightenment, where exploring and documenting the natural world took hold as highly valuable societal goods. But this itch for discovery and the coddiwompleness attendant with it are not necessarily, in all cases, salutary or noble. Nor is it limited to wanderlust and *fernweh*, that “farsickness” for an experience of travel and the unknown. Indeed, Gottfredson and Hirschi, who offered one of the most successful criminological theories to date, describe “adventurousness” as a characteristic of low self-control—the degree to which a person is vulnerable to various forms of antisocial, risky, and dangerous behaviors, especially in the moment^1^.

Various aspects of adventurousness, especially those related to outdoor activities, have been associated with both positive and negative psychological outcomes. Adventurousness, for example, may improve well-being and happiness and is linked with appreciating nature, relaxation, and learning^2^. Yet, in contrast, adventurousness is intertwined with risky behaviors and sensation seeking (a strong need for novelty) that place people in high-risk situations that sometimes lead to serious injury or death^2^. Since non-experimental studies can suffer from confounding and reverse causation, and personality traits cannot be randomized, it is challenging to unravel how adventurousness actually impacts the mind, decision making, and ultimately behavior.

Fortunately, in 2019, a large genome-wide association (GWA) study was performed, using 23andMe data, that found genetic variants (single-nucleotide polymorphisms, SNPs) strongly associated with the self-reported tendency to be “adventurousness” or “cautious”. The genetic summary data for adventurousness (effect estimates and standard errors) can be harnessed and used to study the impact of adventurousness on other traits for which large GWA studies have also been performed. Doing so abates many of the concerns for confounding and reverse causation to which observational designs are prone, supposing certain assumptions hold (see Methods).

To that aim, we use a form of Mendelian randomization (MR) to explore the relationship between adventurousness and 10 cognitive and behavioral traits: neuroticism, education years, ordinary (non-clinical) mood swings, subjective well-being, duration of walking for pleasure, propensity to speed in an automobile, lifetime number of sexual partners, age first had sexual intercourse, fluid intelligence score, and lifetime smoking. We provide a snapshot of how genetically influenced, self-reported adventurousness influences human behavior.

## Results

**Table 1** contains the results for the MR tests of adventurousness on 10 cognitive and behavioral traits. What follows below is a trait-specific report of the results. The inverse-variance weighted (IVW) findings are given, along with a discussion of them in comparison to the MR-Egger, weighted median, and weighted mode estimators (“sensitivity estimators”). The comparison is with regard to the directions and magnitudes of their effects (not their *P*-values) (see Methods).

**Table 1.**
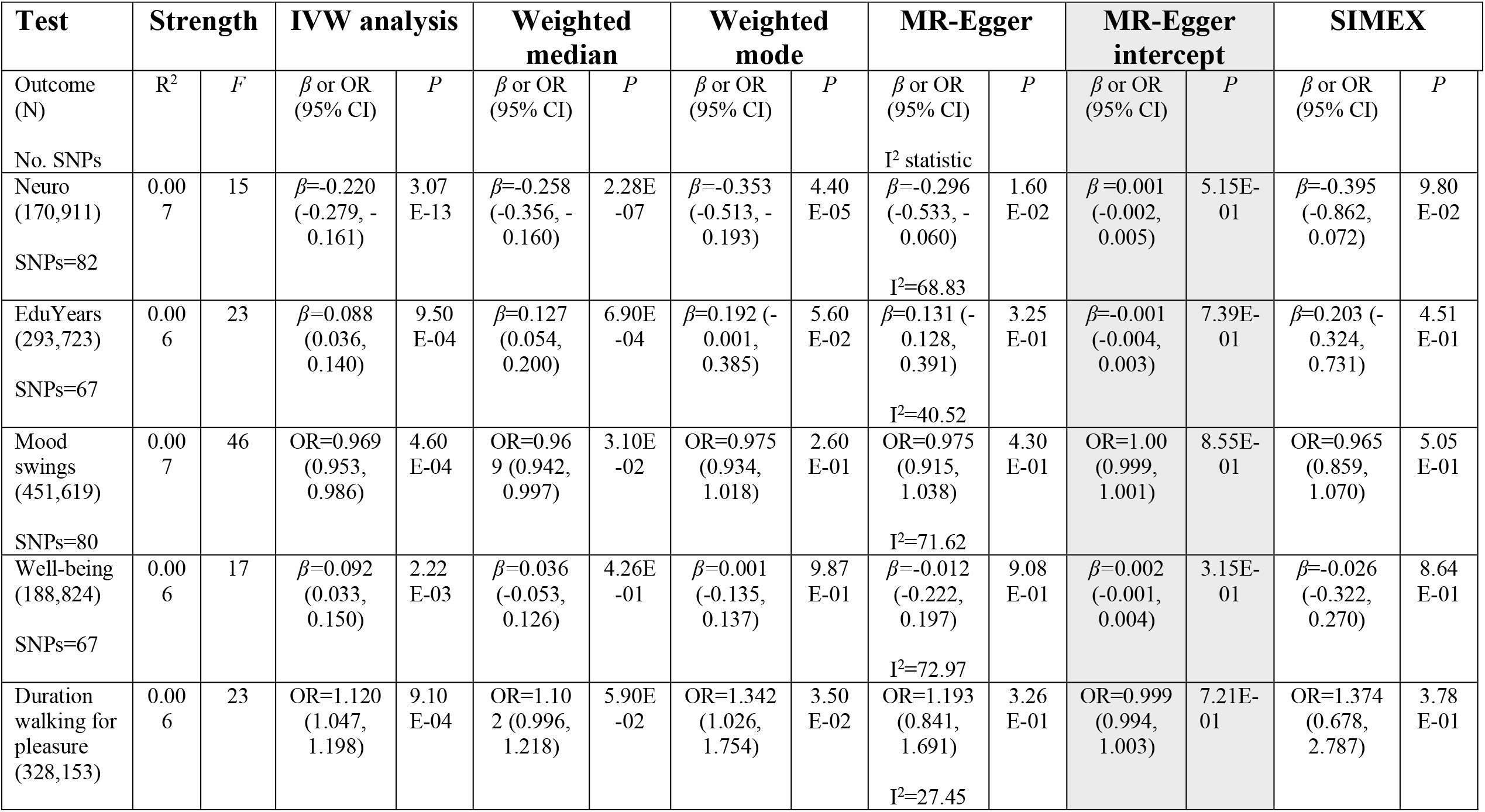
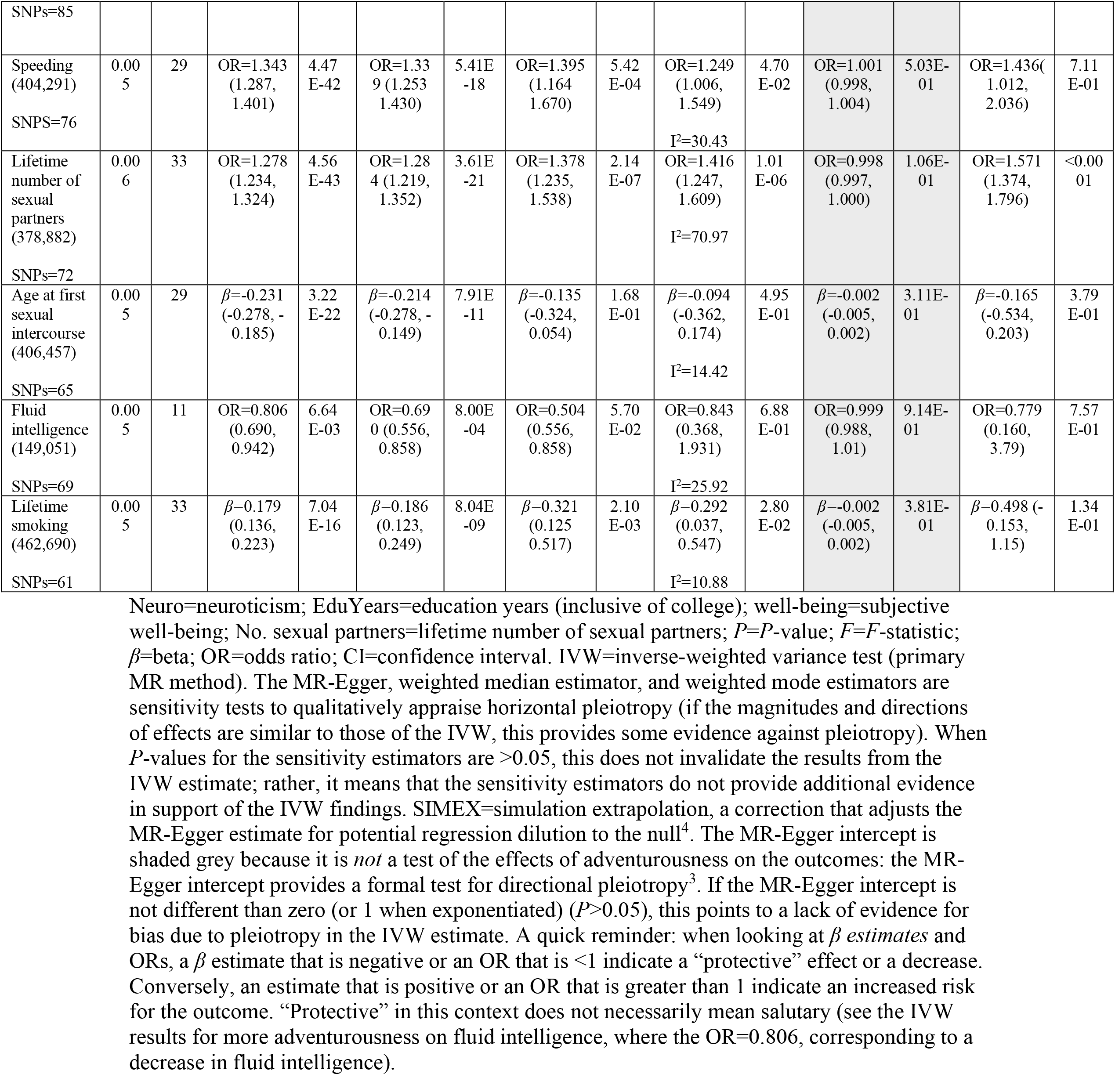
Estimates of the effect of adventurousness on 10 cognitive and behavioral traits.

### Neuroticism

Adventurousness decreased neuroticism: IVW beta estimate (*β*) estimate −0.220; 95% CI −0.279, −0.161; *P*=3.07E-13). The sensitivity estimators aligned in direction and magnitude of effects.

### Education years

Adventurousness increased years of schooling: IVW *β* estimate 0.088; 95% CI 0.036, 0.140; *P*=9.50E-04). The sensitivity estimators aligned in direction and magnitude of effects.

### Mood swings

Adventurousness was slightly protective against mood swings: IVW odds ratio (OR) estimate 0.969; 95% CI 0.953, 0.986; *P*=4.60E-04). The sensitivity estimators aligned in direction and magnitude of effects.

### Subjective well-being

Adventurousness increased subjective well-being: IVW *β* estimate 0.092; 95% CI 0.033, 0.150; *P*=2.22E-03). The sensitivity estimators are discrepant both in direction and magnitude of effects, suggesting bias due to pleiotropy.

### Duration of walking for pleasure

Adventurousness increased the duration of walking for pleasure: IVW OR estimate 1.120; 95% CI 1.047, 1.198; *P*=9.10E-04). The sensitivity estimators mostly aligned in direction and magnitude of effects, with the exception of a larger magnitude of effect observed for the weighted mode estimator (1.342, relative to the IVW’s 1.120). This possibly indicates bias due to pleiotropy.

### Propensity for speeding in an automobile

Adventurousness increased the propensity for speeding in an automobile: IVW OR estimate 1.343; 95% CI 1.287, 1.401; *P*=4.47E-42). The sensitivity estimators aligned in direction and magnitude of effects.

### Lifetime number of sexual partners

Adventurousness increased the lifetime number of sexual partners: OR estimate 1.278; 95% CI 1.234, 1.324; *P*=4.56E-43). The sensitivity estimators aligned in direction and magnitude of effects.

### Age first had intercourse

Adventurousness lowered the age of first having sexual intercourse: IVW *β* estimate −0.231; 95% CI −0.278, −0.185; *P*=3.22E-22). The sensitivity estimators aligned in direction and magnitude of effects.

### Fluid intelligence

Adventurousness lowered fluid intelligence: IVW OR estimate 0.806; 95% CI 0.690, 0.942; *P*=6.64E-03). The sensitivity estimators mostly aligned in direction and magnitude of effects, with the exception of the weighted mode, for which the estimate dipped downwards (0.504, relative to the IVW’s 0.806). This possibly indicates bias due to pleiotropy.

### Lifetime smoking

Adventurousness increased the lifetime smoking: IVW *β* estimate 0.179; 95% CI 0.136, 0.223; *P*=7.04E-16). The sensitivity estimators aligned in direction and magnitude of effects.

### MR-Egger intercept

The MR-Egger intercept column is shaded grey because it is not a test of the associations between adventurousness and the outcome traits. The MR-Egger intercept column reports a formal test for directional pleiotropy and an assessment of the validity of the instrument assumptions^3^. If the intercept is not different than 0 (or 1 on the exponentiated scale), this points to a lack of evidence for bias in the IVW estimate. For all associations, the MR-Egger intercept test demonstrated no evidence for directional pleiotropy (*P*>0.05).

### Simulation extrapolation (SIMEX)

SIMEX is a correction procedure that adjusts the MR-Egger estimate for potential regression dilution to the null^4^. Sometimes a discrepancy between the MR-Egger and other MR estimators can be due to regression dilution, especially if the I^2^ statistic for the MR-Egger estimate indicates that there may be bias in the MR-Egger estimate. When the I^2^ statistic is <90%, correction procedures are recommended. SIMEX correction was applied but did not change the inference for any of the associations in this study. (The SIMEX-corrected MR-Egger column is not being used for informal comparison with the IVW and other estimators.)

## Discussion

In this analysis of adventurousness on 10 cognitive and behavioral traits, there was evidence that adventurousness “cuts both ways”, dovetailing closely with the multifaceted influences observed in the literature.

In particular, the trait had some positive psychological impacts: it decreased neuroticism and mood swings and increased years of schooling. In contrast, adventurousness also predisposed to risky behaviors, including increasing the number of lifetime sexual partners, decreasing the age of first having sexual intercourse, increasing the propensity to speed in an automobile, and increasing lifetime smoking. There was suggestive evidence that adventurousness decreased fluid intelligence and improved both subjective well-being and duration of walking for pleasure, but the sensitivity estimators pointed to evidence for potential pleiotropic bias for these traits. The clearest and strongest evidence for adventurousness on any of the tested traits is for increasing the lifetime number of sexual partners. Not only did the IVW and sensitivity estimators comport, but the MR-Egger estimator also had a *P*-value below the Bonferroni threshold.

That adventurousness increases the years of staying in higher education but possibly decreases fluid intelligence is intriguing, since schooling years and fluid intelligence are bidirectionally causally related (they influence each other)^5–7^. Perhaps the “hit” to fluid intelligence is related to the detrimental effects of the risky behaviors. However, it might also be that the finding for fluid intelligence is biased from pleiotropy and/or that the predominant causal direction is the other way around—i.e., it may be that fluid intelligence influences adventurousness (not that adventurousness influences intelligence).

When the full summary data for the adventurousness GWA study are available (at present only the data for the top findings are public), bidirectional and multivariable analyses including adventurousness and some of these traits could be performed. For instance, it would be interesting to see whether smoking partly mediates the observed decrease in fluid intelligence that seems to be due to adventurousness. If both smoking and adventurousness were included in a multivariable model, this could be assessed. If the estimate for fluid intelligence observed in the univariable model attenuates to the null when accounting for smoking, this would provide evidence that smoking may be the determinantal determinant on fluid intelligence.

The primary strength of this study is that it capitalizes on the power of 11 large GWA studies to provide a picture of how adventurousness impacts psychological phenotypes. It is the most comprehensive MR investigation of adventurousness on the mind to date.

The study has several limitations. All MR studies critically rely on the validity of the instrumental variables. We triangulated the IVW estimate with a battery of sensitivity estimators that, for the most part, provided evidence against violations due to unwanted pleiotropy, except for fluid intelligence, subjective well-being, and duration of walking for pleasure. We also only used instruments lacking between-SNP heterogeneity, reducingconcern for type 1 errors.

A final concern involves the measurement of adventurousness. The 23andMe data utilized a simple (and limited) item to gauge variation on a complex psychological phenotype. This introduces some degree of measurement error and will not fully capture the spectrum of variation for a multi-faceted dimensional trait like that of adventurousness (for a thorough overview of the limitations of short/single item personality measures, see^8^).

Nonetheless, this does not mean that the current results are meaningless, only that additional work is needed to replicate the findings. Moreover, it also suggests that an increased focused on sound psychometrics may be worthwhile and benefit both the large direct-to-consumer genomic consortia, as well as the research community as a whole. Consumers would receive more valid and reliable insight, and researchers would be able to access data with less measurement error and more desirable psychometric and research qualities overall^8^. As is, the simple construct of self-reported adventurousness evinced bivalent influences and underscored the reality that trade-offs often accompany many human personality constructs.

Our findings set the stage for some interesting research which will help to further illuminate how personality variation shapes the types of lives and the nature of experiences that people encounter throughout their lives.

## Methods

### Conceptual approach

Although not exactly (there are some important differences), heuristically, MR is analogous to a randomized controlled trial (RCT). In an RCT, treatment allocation is randomized by investigator. With MR, quasi-randomization occurs due to alleles randomly assorting from parent to offspring. MR exploits this and two other features of the genome (genotype assignment at conception and pleiotropy, genes influencing more than one trait^9–11^), in an instrumental-variables framework.

MR largely averts two serious epidemiologic problems that arise from the inability to randomize: confounding and reverse causation. Using SNPs instrumentally avoids most environmental sources of confounding, and the fixed nature of genotype assignment at conception avoids most sources of reverse causation.

Two-sample MR is an adaptation of the procedure that uses summary statistics from two GWA studies^3,12–16^. Typically, with two-sample MR, the IVW method is the standard approach (Fig. 1 contains an example).

**Fig. 1.**
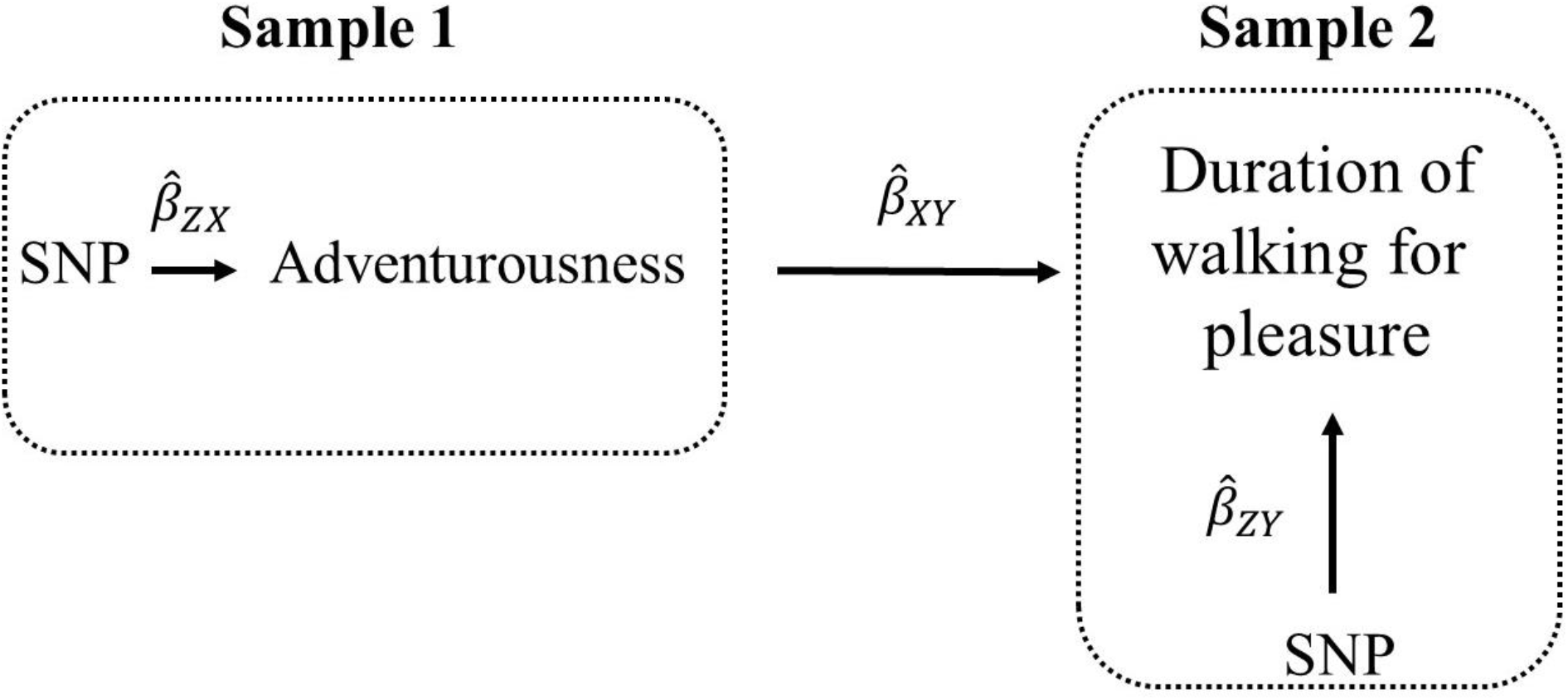
Illustration of two-sample MR using the test of the causal effect of adventurousness on the duration of walking for pleasure as an example. Estimates of the SNP-adventurousness associations 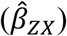 are calculated in sample 1 (from a GWA study of adventurousness). The association between these same SNPs and the duration of walking for pleasure are then estimated in sample 2 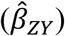 (from a GWA study of the duration of walking for pleasure). For each instrumental SNP (chosen for being associated at genome-wide significance with adventurousness and not in linkage disequilibrium with other SNPs strongly associated with adventurousness), 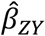 is the change in log odds for duration of walking for pleasure per copy of the effect allele, and 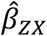 is the log odds change in adventurousness per copy of the effect allele. The estimates from the instrumental SNPs are combined into Wald ratios 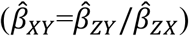 The 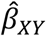 ratio estimates are meta-analyzed using the inverse-variance weighted (IVW) analysis 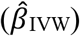 method. The IVW method is the standard approach for MR with two-sample summary data. In this example, it produces an overall causal estimate of adventurousness on the duration of walking for pleasure. The IVW method assumes all the multi-allelic SNPs for adventurousness are valid. If any of the instrumental SNPs are invalid due to a violation of the instrumental variable assumptions, this can lead to bias in the causal estimate^17^. Sensitivity estimators are used to judge whether the causal estimate is plausible (more below).

### Mendelian randomization assumptions

MR relies on the validity of three assumptions^18^. In the context of the present analysis, these assumptions are as follows: (i) the SNPs acting as the instrumental variables for adventurousness are strongly associated with adventurousness; (ii) the adventurousness SNPs are independent of confounders of adventurousness and the outcomes of interest; and (iii) the adventurousness SNPs are associated with the outcomes of interest only through adventurousness (no horizontal pleiotropy; the SNPs are not associated with the outcomes independent of adventurousness^14,18^).

### Adventurousness (instrument) data source

Linnér *et al*. (2019)^19^ performed a GWA study of adventurousness (defined as the self-reported tendency to be “adventurous” versus “cautious”) in research participants from 23andMe (n=557,923) with European ancestry. Adventurousness, an ordered categorical variable, was analyzed from responses to the question “If forced to choose, would you consider yourself to be more cautious or more adventurous?”, with possible responses ranging from “(1) very cautious” to “(5) very adventurous”. The GWA study was adjusted for a minimum of 10 principal components, sex, and birth years.

Estimates of the proportion of variance in adventurousness explained by the genetic instruments (R^2^) and the strength of the association between the genetic instruments and adventurousness (*F*-statistics) were generated for each of the tests. *F*-statistics can be used to examine whether results suffer from reduced statistical power to reject the null hypothesis. This can happen if the adventurousness instruments explain a limited proportion of the variance in adventurousness. *F*-statistics <10 are conventionally considered to be weak^20,21^. The *F*-statistics and R^2^ values used to calculate them are presented in Table 1.

### Selection of outcome data sources

MR-Base (available at http://app.mrbase.org/)^12^, a public collection of GWA studies with summary statistics for use in MR studies, was used to identify cognitive and behavioral outcome traits with available GWA study summary data. Only studies that contained more than 100,000 subjects were considered in order to maximize the power of two-sample MR. Eight traits available through MR-Base were selected for examination: neuroticism, education years, mood swings, well-being, duration of walking for pleasure, lifetime number of sexual partners, age first had intercourse, and fluid intelligence. In addition, two GWA studies not available in MR-Base but publicly available were also included: propensity for speeding in an automobile^19^ and lifetime smoking^22^.

### Neuroticism

As part of the Social Science Genetic Association Consortium (SSGAC), Okbay *et al*. (2016)^23^ performed a GWA study of neuroticism in research participants of European ancestry within the Genetics of Personality Consortium (GPC) and UK Biobank (n=170,911)^24,25^. The neuroticism measure for the UK Biobank participants came from their score on a 12-item version of the Eysenck Personality Inventory Neuroticism scale^23,26^. The neuroticism batteries used by the GPC cohorts varied but were harmonized. The meta-analyzed GWA studies adjusted for varying numbers of principal components, sex, and age. The summary data are reported in standard deviation (SD) units.

### Education years

As part of the SSGAC, Okbay *et al*. (2016) performed a GWA study of education years in participants of European ancestry (n=293,723)^27^. Education years was measured for those who were at least 30 years of age, and International Standard Classification of Education (ISCED) categories were used to impute a years-of-education equivalent (SNP coefficients per SD units of years of schooling). The GWA study was adjusted for 10 principal components, age, and sex^27^. The summary data are reported in standard deviation (SD) units.

### Mood swings

The UK Biobank appraised ordinary (non-clinical) ups and downs (mood swings) with the question “Does your mood often go up and down?” (UK Biobank data-field 1920). Members of the Medical Research Council-Integrative Epidemiology Unit (MRC-IEU) at the University of Bristol used PHESANT^28^ to run a GWA study of this mood-swing-measure (n=204,412 cases; n=247,207 controls). They treated the variable as binary and adjusted for age at recruitment and sex.

### Subjective well-being

As part of the SSGAC, Okbay *et al*. (2016)^23^ performed a meta-analyzed GWA study of subjective well-being in research participants of European ancestry (n=197,174), excluding individuals from 23andMe. Subjective well-being was broadly defined to include positive and negative subjective evaluations and the analysis pooled both “positive affect” and “life satisfaction” measures, harmonized across 59 participating cohorts. The meta-analyzed GWA studies adjusted for varying numbers of principal components, sex, and age. The summary data are reported in standard deviation (SD) units.

### Duration of walking for pleasure

The UK Biobank appraised duration of walking for pleasure with the question “Each time you went walking for pleasure, about how long did you spend doing it” (UK Biobank data-field 981). Members of the MRC-IEU used PHESANT^28^ to run a GWA study of this variable for duration of walking for pleasure (n=328,153). They treated the variable as an ordered categorical type and adjusted for age at recruitment and sex.

### Propensity for speeding in an automobile

Linnér *et al*. (2019)^19^ performed a GWA study of the propensity to speed in an automobile in participants in the UK Biobank (n=404,291) with European ancestry. Respondents were asked, “How often do you drive faster than the speed limit on the motorway?” Response options included: 1) Never/rarely; 2) Sometimes; 3) Often; 4) Most of the time; and 5) Do not drive on the motorway. The GWA study was adjusted for a minimum of 10 principal components, sex, and birth years.

### Lifetime number of sexual partners

The UK Biobank asked participants how many sexual partners they had had in their lifetimes (UK Biobank data-field 2149). Members of the MRC-IEU used PHESANT^28^ to run a GWA study of this variable for lifetime number of sexual partners (n=378,882). They treated the variable as an ordered categorical type and adjusted for age at recruitment and sex.

### Age first had intercourse

The UK Biobank asked participants at what age they first had sexual intercourse (defined as including vaginal, oral or anal intercourse) (UK Biobank data-field 2139). Members of the MRC-IEU used PHESANT^28^ to run a GWA study of this variable for age of first having sexual intercourse (n= 406,457). They treated the variable as continuous and adjusted for age at recruitment and sex.

### Fluid intelligence

The UK Biobank appraised fluid intelligence by summing the number of correct answers given to 13 fluid intelligence questions (UK Biobank data-field 20016). Members of the MRC-IEU at the University of Bristol used PHESANT to run a GWA study of this fluid intelligence measure (n=149,051)^28^. They treated the variable as an ordered categorical type and adjusted for age at recruitment and sex.

### Lifetime smoking

Wooten *et al*. (2018) performed a GWA study of lifetime smoking, which adjusted for sex and genotyping chip, among participants in the UK Biobank (n=462,690)^22^. This novel measure of smoking is inclusive of smoking status, smoking duration, heaviness, and cessation: a standard deviation (SD) increase in lifetime smoking is “equivalent to an individual smoking 20 cigarettes a day for 15 years and stopping 17 years ago or an individual smoking 60 cigarettes a day for 13 years and stopping 22 years ago”^22^. Standardized betas were calculated by dividing both betas and standard errors by the SD of lifetime smoking in the whole sample (SD=0.694). The full summary data is publicly available at https://data.bris.ac.uk/data/dataset/10i96zb8gm0j81yz0q6ztei23d.

### Instrument construction

For the adventurousness instruments (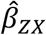 in Fig. 1), SNPs associated at genome-wide significance (*P* < 5 × 10^−8^) with adventurousness were extracted from the Linnér *et al*. (2019)^19^ GWA study of adventurousness. The SNPs were independent (not in linkage disequilibrium, LD, with an r^2^ < 0.001, at a clumping distance of 10,000 kilobases with reference to the 1000 Genomes Project (http://www.internationalgenome.org/). The corresponding effect estimates and standard errors for these SNPs were then obtained from the outcome GWA studies (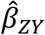 in Fig. 1).

When a SNP was not available in the outcome datasets, a “proxy” SNP in LD with the SNP at r^2^≥0.80 (assessed using 1000 Genomes Project) was chosen. If the “proxy” SNP was not available in the outcome dataset, the SNP was removed from the analysis. SNP-exposure and SNP-outcome associations were harmonized with the “harmonization_data” function within the MR-Base “TwoSampleMR” package within R^12,29^. Harmonized SNP-exposure and SNP-outcome associations were combined with the IVW method (Fig. 1).

For all tests, RadialMR regression^30^ was run to detect SNP outliers. Outlier SNPs were removed. (A different number of adventurousness SNPs were used for the various tests due to outliers being removed and whether a SNP or its “proxy” was available in the outcome dataset.) All instrumental variables included in this analysis have Cochrane’s *Q*-statistic *P*-values indicating no evidence for heterogeneity between SNPs^31^ (heterogeneity statistics are provided in Supplementary Tables 11-20).

### Sensitivity analyses

A liability of the primary IVW estimator is that its estimate can be biased if one or more the SNPs in its multi-allelic instrument are directionally pleiotropic (where the horizontally pleiotropic effect does not average to zero)^32^. To assess possible violations to MR assumption (iii)—about horizontal pleiotropy—MR-Egger regression, weighted median, and weighted mode MR methods were run and their results triangulated with those of the IVW.

Triangulation is a qualitative method that integrates knowledge from approaches with different assumptions^33^. In this case, the various MR estimators make different assumptions about the underlying nature of pleiotropy. When the magnitudes and directions of the various MR methods comport across estimators, this relative homogeneity is a qualitative screen against pleiotropy that makes causation more plausible^34^. Substantial violations to the MR assumption about no pleiotropy make it unlikely there would be homogeneity in the direction and magnitudes of their effects.

Thorough descriptions of the various MR methods and the different assumptions they make about pleiotropy are described elsewhere^32,35,36^. However, briefly, unlike the IVW, the MR-Egger sensitivity estimator can provide unbiased estimates of causal effects, supposing all instrumented SNPs are invalid due to pleiotropy, conditional on two additional assumptions holding: 1) the potential pleiotropic effects of the instrumented SNPs are independent of their strength and 2) measurement error in the genetic instrument being negligible. The weighted median estimator can provide unbiased causal effects, assuming at least 50% of the chosen SNPs are valid. The weighted mode estimator assumes the most common effect estimate among SNPs in an instrument comes from a valid instrument.

In addition to the comparative sensitivity estimators, a SIMEX correction was performed to correct potentional regression to the null in the MR-Egger estimates^37^.

### Number of tests

In total, 10 MR tests were run (detailed characteristics for the individual SNPs used in each model are provided in Supplementary Tables 1-10). To account for multiple testing across analyses, a Bonferroni correction was used to establish a *P*-value threshold for strong evidence (*P* < 0.005) (false-positive rate = 0.05/10 outcomes).

### Statistical software

SIMEX corrections were perfomed in Stata SE/16.0^38^. All other described analyses were performed in R version 3.5.2 with the “TwoSampleMR” package^12^.

## Data Availability

All data sources used for SNP-exposure and SNP-outcome associations are publicly available. The summary data for the adventurousness instruments and propensity for speeding in an automobile are available at https://www.thessgac.org/data. The full lifetime-smoking summary data are available at https://data.bris.ac.uk/data/dataset/10i96zb8gm0j81yz0q6ztei23d. The remaining data used for these analyses are accessible within MR-Base: http://www.mrbase.org/.

## Data availability

All data sources used for SNP-exposure and SNP-outcome associations are publicly available. The summary data for the adventurousness instruments and propensity for speeding in an automobile are available at https://www.thessgac.org/data. The full lifetime-smoking summary data are available at https://data.bris.ac.uk/data/dataset/10i96zb8gm0j81yz0q6ztei23d. The remaining data used for these analyses are accessible within MR-Base: http://www.mrbase.org/^12^.

## Acknowledgements

Thank you to the cohorts that made their GWA study summary data public.

## Additional information

Supplementary information accompanies this paper.

## Footnotes

1 The original source of the quote appears lost, but David Brooks attributes the quote to George Eliot in his book *The Road to Character*^39^.

